# How will this continue? Modelling interactions between the COVID-19 pandemic and policy responses

**DOI:** 10.1101/2020.03.30.20047597

**Authors:** Axel G. Rossberg, Robert J. Knell

**Affiliations:** School of Biological and Chemical Sciences, Queen Mary University of London, Mile End Road, London E14NS

## Abstract

Much of the uncertainty about the progression of the COVID-19 pandemic stems from questions about when and how non-pharmaceutical interventions (NPI) by governments, in particular social distancing measures, are implemented, to what extent the population complies with these measures, and how compliance changes through time. Further uncertainty comes from a lack of knowledge of the potential effects of removing interventions once the epidemic is declining. By combining an epidemiological model of COVID-19 for the United Kingdom with simple sub-models for these societal processes, this study aims to shed light on the conceivable trajectories that the pandemic might follow over the next 1.5 years. We show strong improvements in outcomes if governments review NPI more frequently whereas, in comparison, the stability of compliance has surprisingly small effects on cumulative mortality. Assuming that mortality does considerably increase once a country’s hospital capacity is breached, we show that the inherent randomness of societal processes can lead to a wide range of possible outcomes, both in terms of disease dynamics and mortality, even when the principles according to which policy and population operate are identical.. Our model is easily modified to take other aspects of the socio-pandemic interaction into account.

## Introduction

The use of detailed simulation modelling to assess the potential outcomes of the current SARS-CoV-2 pandemic and its associated disease, COVID-19, has produced crucial information for policy makers and healthcare managers, most notably in the UK where a detailed modelling study (Ferguson *et al*. 2020) has led to a significant change in government policy regarding the pandemic (Gallagher 2020). The model used, an adaptation of one that was previously used to model potential influenza pandemics (Ferguson *et al*. 2006), is a complex individual-based simulation with many thousand lines of code which are not yet available openly (Tweet from @neil_ferguson 22nd March 2020). While such models can make accurate predictions for given assumptions on individual behaviour, their computationally intensive nature means that they do not easily capture a major source of uncertainty determining disease dynamics: when and how non-pharmaceutical interventions (NPI) such as social distancing measures are enacted by governments and how the population change their behaviour as a result. To address this aspect of the problem, we propose to use simpler models and perform “Management Strategy Evaluation” (MSE: (Bunnefeld, Hoshino & Milner-Gulland 2011; Punt *et al*. 2016), an adaptive framework for comparing multiple management strategies in a variety of circumstances. Originally developed for assessing fisheries management schemes, this framework has the potential to be applied widely for assessing best practice in other important management scenarios (Bunnefeld *et al*. 2011).

Using a model based on the “Susceptible - Infected - Recovered” (SIR) framework with the infected population divided into infectious and noninfectious, and symptomatic and asymptomatic compartments to simulate SARS-CoV-2 in the UK population, we explore the potential effects of two important aspects of pandemic management: the frequency at which of management interventions are reviewed, and the potential decline in population compliance with NPI. Responses to COVID-19 have varied widely between nations in both the speed and the stringency of the interventions used (Hale & Webster 2020). South Korea, for example, intervened very quickly once the first cases were reported whereas the response has been considerably slower in countries such as the UK and the USA (Fig. 1). It is likely that the continuing response to the pandemic will also vary between nations, with different governments choosing to relax the stringency of their interventions at different stages of epidemic decline. The importance of early intervention at the start of a pandemic to reduce mortality is well known, but there has been little assessment to date of the effects of removing management interventions at different points once the epidemic in a particular nation or area has started to decline.

**Figure 1.**
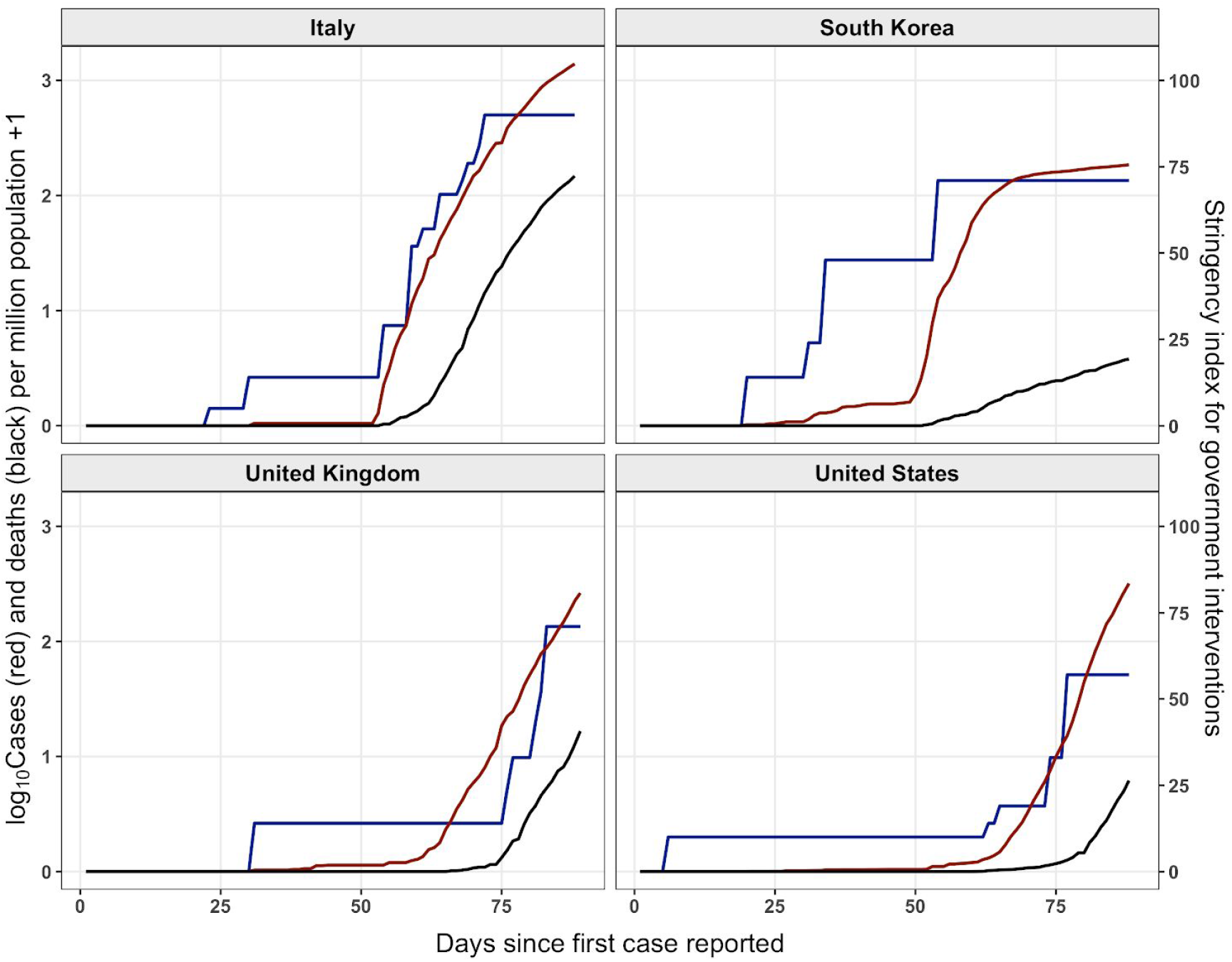
Government intervention stringency (blue) and the log_10_ numbers of reported cases (red) and deaths (black) per million people plotted against the days since the first reported case for four nations. Data from the Oxford COVID-19 Government Response Tracker (Hale & Webster 2020), downloaded 30th March 2020.

Many of the interventions that are necessary to reduce the transmission of a dangerous infectious agent spread by close contact involve restricting the activities of the population in order to reduce contact rates. Further interventions seek to change behaviour to reduce the probability of infection per contact by, for example, emphasising the importance of frequent hand-washing and by isolating potentially infectious cases. Compliance with these interventions will vary between individuals depending on a wide variety of factors, and because of the restrictive nature of the interventions it is likely that compliance will decay over time, especially if the public perception is that the risk is low or the epidemic is declining. This decline in compliance has been found in a number of studies of public behaviour, notably during recent influenza epidemics in Hong Kong (Liao *et al*. 2019), Malaysia (Wong & Sam 2010) and the Netherlands (de Zwart *et al*. 2010). Despite the importance of behavioural changes to the progress of epidemics (Ferguson 2007; Bauch & Galvani 2013; Yan, Tang & Xiao 2018), behavioural changes over time are rarely incorporated into epidemic models (Funk *et al*. 2015). The factors determining how an individual’s behaviour changes with respect to an epidemic threat will include the degree of perceived risk and the perceived benefits from a particular behavioural change (Yan *et al*. 2018), cultural and religious beliefs, expectations and norms and the interaction between an individual and both traditional news media and social media influences. These will all vary between nations and so the degree of initial compliance and the rate of decline of compliance over time will vary and thus it is important to explore the potential impacts of this heterogeneity on the development of the epidemic.

## Methods

### Structure of epidemiological model

We model disease dynamics using a variant of the conventional SIR model that distinguishes the *Susceptible* (S), *Infected* (I), and *Recovered* (R) stages of disease progression in individuals. In order to model inherent delays and observational constraints, we distinguish amongst infected individuals in addition amongst those that are symptomatic (label: s) and asymptomatic (label: a), and in both cases between the early stages that do not (label 1) and late stages that do (label 2) transmit the disease. Thus infected individuals belong to one of the four classes Ia1, Ia2, Is1, and Is2. We include in the R stage both immune individuals and fatalities. We simulate disease dynamics in discrete, 24h time steps, applying methods of matrix population dynamics (Caswell 2006). Disease mortality is computed using a separate model (see below). We assume a total initial total population size of *N* = 66·10^6^, representative of the United Kingdom (UK). We seeded the model with 10 asymptomatic early stage individuals, i.e., we start with *S* = *N* - 10, *I*_s1_ = 10, *I*_a1_ = 10, *I*_s2_ = 0, *I*_a2_ = 0, using Italic symbols to represent population numbers in corresponding stages.

### Parameterization of disease dynamics

The mean time to transition from Ia1 to Ia2 and from Is1 to Is2 is taken as 4.6 days (Ferguson *et al*. 2020). Also following Ferguson *et al*. (2020), a proportion of 20% of infected individuals are assumed to become symptomatic (Is1, Is2), and the symptomatic individuals in stage 2 are assumed to be twice as infectious as the asymptomatic ones. The relation between the basic reproduction number R_0_ and the linear growth rate *r*_0_ (or corresponding doubling time) of a disease is known to depend sensitivity on the statistical distribution of the time intervals between transmissions, which our model does not represent well. We therefore compute R_0_indirectly by first determining *r*_0_ from our model and then computing R_0_ = (1 + *r*_0_ *T*_gen_ / α)^α^, where *T*_gen_ = 6.5 days is the mean generation time (i.e. time between transmission) following Ferguson et al. (2020) and α =2 is the shape parameter of an assumed gamma distribution of time between transmissions (Park *et al*. 2019). Solving for *r*_0_, we obtain *r*_0_ = α (R_0_ ^1/α^ - 1) / *T*_gen_ = 0.17 day^-1^, implying a doubling time of about 4 days.

New infection arise at a rates are proportional to the product (*I*_s2_ + 0.5 *I*_a2_) × *S* / *N*. The corresponding proportionality constant, infection rate, and the rate for recovery, i.e. of transitions from stages Ia2 and Is2 to R, were determined such that the model’s intrinsic values for mean generation time *T*_gen_ and linear growth rate *r*_0_ match those above. To obtain these values we first constructed the population projection matrix **L** representing the processes above for low disease prevalence (*S* → *N*) and determined its dominating eigenvalue λ and the corresponding right and left eigenvectors, **w** and **v**, respectively. We then decomposed **L** = **F** + **S**, where **F** (“fertility matrix”) represents transitions between stages involving new infections and **S** (“survival matrix”) all other transitions. We then computed *r*_0_ (in units of day^-1^) as ln(λ) and *T*_gen_ (in days) as 1 + (**v**^T^**Sw**)/(**v**^T^**Fw**) (Bienvenu & Legendre 2015). The fitted value for infection rate as defined above is 1.026 day^-1^, and for the recovery rate 0.149 day^-1^. From the policy perspective, the linear growth rate *r*_0_ of the disease and the delays implied by the mean generation time *T*_gen_ are crucial parameters. Using this procedure, we ensure our model reproduces them well.

Denoting by **n**_*t*_ the vector (*S*_*t*_, *I*_a1,*t*_, *I*_a2,*t*_, *I*_s1,*t*_, *I*_s2,*t*_, *R*_*t*_) of the number of individuals in each of the six stages at day *t*, the non-linear dynamics of the model without NPI or pharmaceutical interventions (such as vaccination) is then given by

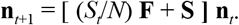

### Mortality

A major policy concern is that at the height of a disease outbreak hospitals will be overloaded, implying inadequate treatment of critical care patients and thus a significantly increased case fatality rate. Current estimates for case fatality for COVID-19 vary widely between countries (Oke & Heneghan), which might partially be attributable to this effect. To model the effect of hospital overload on mortality, we assume that 4.4% of cases will be hospitalized (Ferguson *et al*. 2020), that 30% of hospitalized cases require critical care (Ferguson *et al*. 2020), including ventilators. The number of required ventilators is then computed as the resulting number of cases requiring ventilators, times the ratio between mean duration of critical care — 10 days (Ferguson *et al*. 2020)— and mean duration of infections, which equals mean incubation time plus the inverse recovery rate. We take the number of available ventilators to be 8175 (Parmenter 2020). For the proportion of cases below or reaching critical care capacity we assume a case fatality rate of 0.9%, for cases exceeding hospital capacity a case fatality rate of 9%. Based on these considerations, we compute an effective case fatality rate CFR as a weighted average of these two figures and obtain the number of deaths at a given day in the model as the increment of *R* for this day multiplied by effective CFR.

### Policy responses

The effect of most NPI can be modelled as a reduction in infection rates. We therefore parameterize the effect of NPI by the time-dependent proportion *d*_*t*_ by which infection rates are reduced as a result of NPI (“distancing”). Disease dynamics with policy intervention is thus modeled as

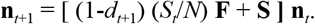

The value of *d*_*t*_ changes from day to day due to two effects: potential policy interventions imposing or relaxing NPI, and variation of compliance through time. We describe the effect of our sub-model for policy interventions in terms of intermediate values *d*_*t*+1/2_, which are then modified in our model for compliance change to compute *dt+*1.

Figure 1 suggests that when COVID-19 breaks out, new policy measures are introduced by governments at irregular intervals, about once every three days in countries like Italy and the UK. In simulations, we therefore sampled each day a random number *x* uniformly distributed between 0 and 1 [notation *x* ∼ Unif(0, 1)], and allowed *d*_*t+*½_ to differ from the previous value *d*_*t*_ only if x < *p*, with *p* = ⅓. In this case, we modelled the decision process leading to potential adjustments of NPI by the following two rules:

#### Rule 1 (imposition of measures)

If the number of “cases” per 1 million individuals, 10^6^ × *I*_s2,*t*_ / *N*_*t*_ is above a threshold of 200 and the daily rate of changes of cases in the last step *r* = ln(*I*_s2,*t*_, *I*_s2,*t*-1_) is positive, then (1 - *d*_*t+*½_) = (1 - *d*_*t*_) *f* [i.e. *d*_*t+*½_ = 1 - (1 - *d*_*t*_) *f*], with *f* ∼ Unif(0.5, 1).

#### Rule 2 (relaxation of measures)

If 10^6^ × *I*_s2,*t*_ / *N*_*t*_ is below the threshold of 20 and the daily rate of change *r* is below −0.1 (indicating a robust decline), then (1 - *d*_*t+*½_) = min[1, (1 - *d*_*t*_) *f*], with *f* ∼ Unif(1, 1.5).

The threshold of 200 for 10^6^ × *I*_s2,*t*_ / *N*_*t*_ used in Rule 1 is motivated by the data for Italy, the United Kingdom, and the USA seen in Fig. 1, and the assumption that each confirmed case in this data corresponds to about 20 cases in total. The tenfold lower threshold in Rule 2 assumes extra caution when measures are relaxed. The range (0.5, 1) of *f* values in Rule 1 is motivated by effects of individuals NPI on disease dynamics computed by (Ferguson *et al*. under the assumption of full compliance and the consideration that in reality compliance with new measures and their effectiveness are difficult to predict. The range (1, 1.5) of *f* values in Rule 2 is similarly motivated, in particular by the intrinsic unpredictability of behavioural change, and in addition by the assumption that after relaxation of NPI people will not fully revert to their pre-NPI behaviour.

### Changes in compliance

The literature indicates that overall compliance with NPI can decline through time but that changes are complex and difficult to predict. Besides, one should not assume compliance behaviour to decay uniformly throughout society since some sections of society will cease complying more easily and faster than others. To represent these effects and their intrinsic uncertainty, we superimpose the immediate effects of policy responses on *d*_*t*_ with a random walk with drift for ln(*d*_*t*_) with dynamics that slow down as *d*_*t*_ approaches zero. Specifically we set *d*_*t+*1_ = min[1, *d*_*t+*½_ exp(- *d*_*t+*½_ *δ*)], where *δ* is a normally distributed random number with mean μ = 1/120 and standard deviation σ = 1/30. This parameterization implies substantial compliance decline over a six-months period, but highly unpredictable change in compliance on the time scale of 1-2 months. We also consider the effect of varying the speed of compliance decline μ.

### Model runs

All model runs were simulations over a 1.5 year period. At the end of each run, we computed the total mortality as the proportion of the initial population died according to our model for mortality above, and the proportion that recovered (excluding deceased). Simulation results were placed on a date axis by matching the day where 55 cumulative fatalities have occured with the corresponding date in the UK, 16 March 2020.

## Results

We stress that, given the simplicity of the model and the high uncertainty in its parameterization, quantitative model outputs should not be taken literally. More relevant are the overall trends and patterns that the model predicts.

Simulating the model without policy intervention, we obtain outbreak dynamics similar to those predicted by Ferguson et al. (2020) for this case (Fig. 2), with a peak in May and a decline to half peak infection rates after about half a month.

**Figure 2:**
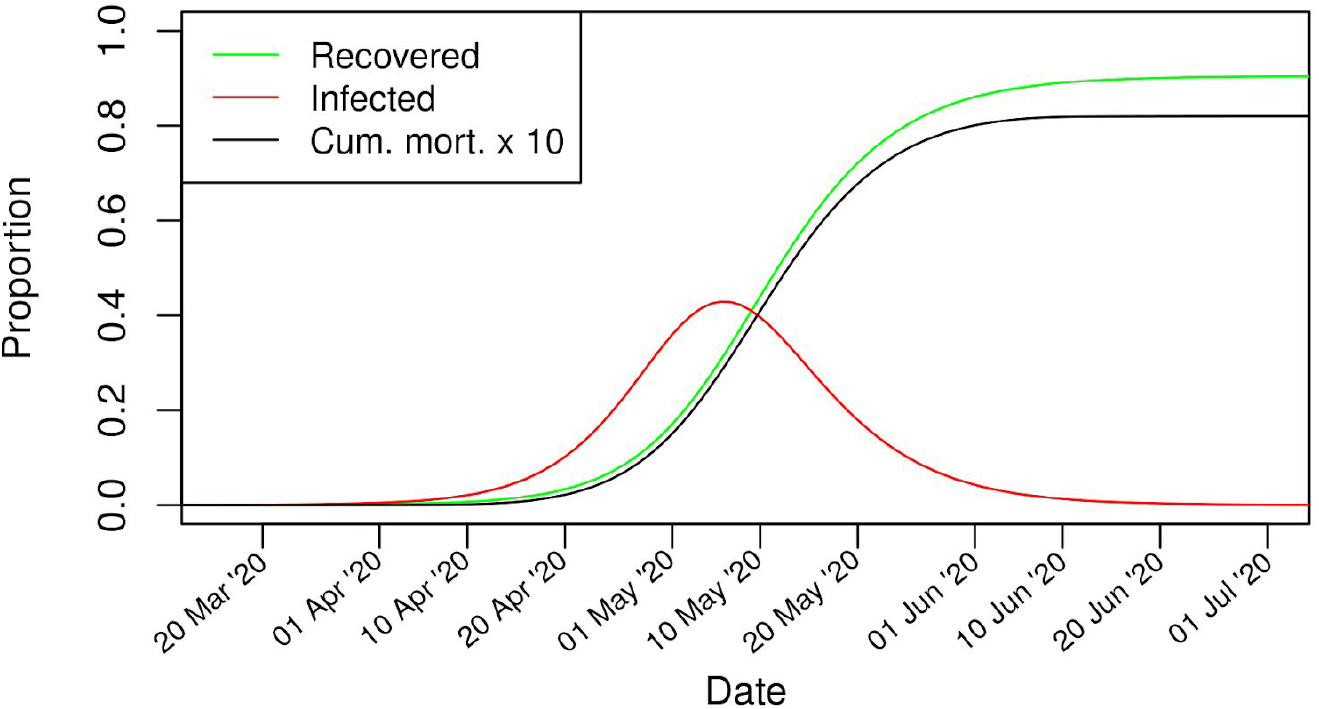
Model simulation without policy responses. The recovered portion here and in figures below excludes the deceased.

The simulation results with policy responses and changing compliance are highly variable. In Fig. 3 we give a few examples of simulated courses of the pandemic. Both large initial outbreaks with high total mortality and less intense outbreaks are possible. In the majority of cases population immunization after the first outbreak remains insufficient for the infections to decline without NPI.

**Figure 3:**
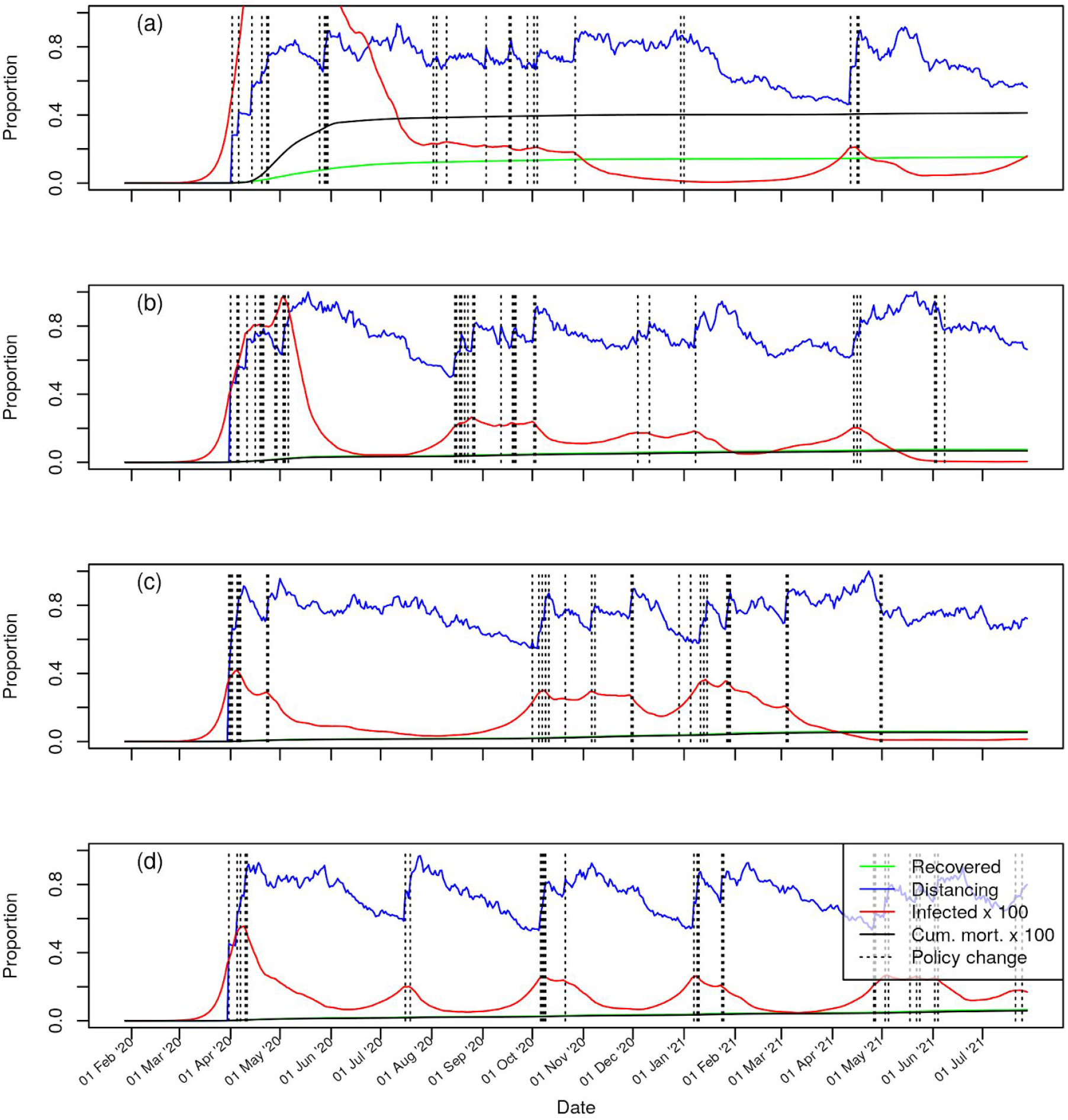
Representative examples of the course of the pandemic with policy responses and declining compliance (standard parameters). Distancing is quantified by the coefficient d_t_ introduced in methods. A slow policy response and high mortality (a); Moderate to fast policy response with irregular secondary outbreaks (b), (c); Fast policy response with regular small secondary outbreaks (d). Standard parameters, simulations differ only by choice of random seed.

To quantify the stress on the health-care system, we define the final mortality rate *m* as the ratio between total mortality and population. This value has a highly skew distribution across multiple model runs. We find *m* > 0.01 indicating significant breaches of hospital capacity, to occur in 29% of runs. In 8.5% of runs we find *m* > 0.03, and in 1% of cases *m* > 0.06. Fatality numbers are similarly skewed across runs. Median total mortality is 0.071%, corresponding to about 47,000 victims of COVID-19. In the lower quarter of runs there are fewer than 37,000 victims but in the upper quarter there are more than 78,000. In 1% of cases there are over 1.7 million deaths.

Next to the uncertainty resulting from the complex interactions between disease, policy and public response described by our model, there is also substantial uncertainty in relevant model parameters. Our model permits us to study these dependencies. For example, if compliance declines not on a time scale of six months but on one of two months (μ = 1/60 in the compliance sub-model), median fatalities nearly double to a value of 91,000, even though overall stress on hospitals does not increase much (*m* > 0.03 in 9.4% of runs). Overall, however, system behaviour is similar to the examples shown in Fig 2, except that policy changes tend to be more frequent. The system is rather robust to declining compliance, because new policy measures are rapidly put in place once a secondary outbreak commences.

We contrast this result with a reduction of the frequency at which NPI are reconsidered, in other words, the speed at which policy responses are enacted. If we modify the original parameterization by halving the rate of policy revisions to one in every six days on average (*p* = ⅙ in the policy sub-model), the median number of fatalities increases more than ten-fold to 522,000. The lower quartile range is then 109,000, the upper quartile range 1.9 million. Hospitals are in most cases overloaded (*m* > 0.03 in 53% of runs). Representative examples for the simulated courses of the pandemic in this case are shown in Fig. 4. In some cases, where the policy response is particularly inefficient, herd immunity is achieved (Fig. 4a), although this rests on the model assumption that immunity is persistent and strong after recovery, something which is not currently known (see Prompetchara et al. 2020 for a review of current knowledge of immune responses to SARS-CoV-2). This herd immunity can lead to removal of all or most NPI after the outbreak. In other cases, case numbers remain high for a long time (Fig. 4d). There can be long time periods without new policy interventions (Figs. 4b,d) but also scenarios with more frequent interventions.

**Figure 4:**
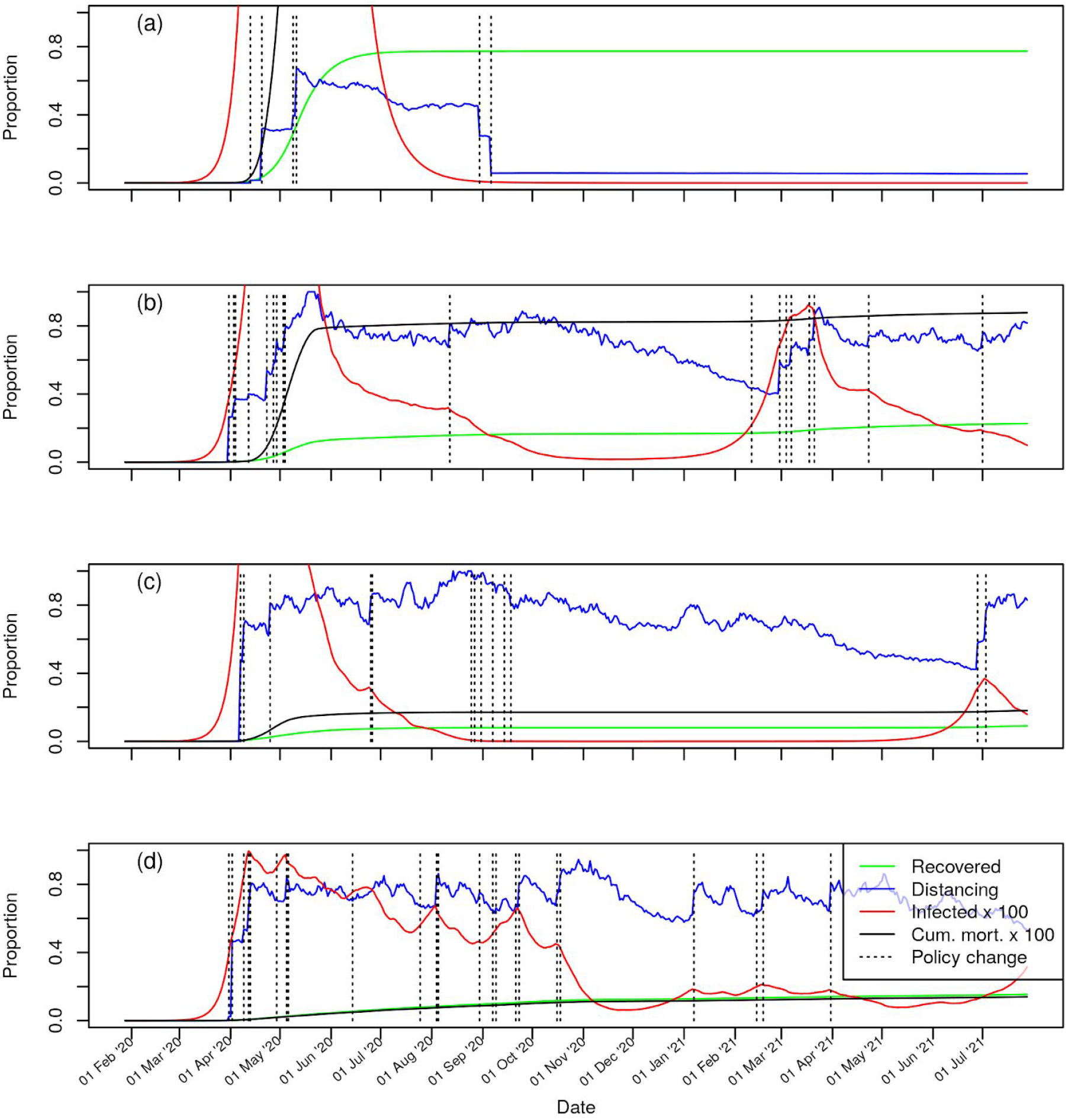
Representative examples of course of pandemic with policy responses and declining compliance (policy revision rate p=1/6), simulations differ only by choice of random seed. With a relatively slow policy response, herd immunity is unintentionally achieved (a), but other types of scenarios can arise with this parameterization as well (b-d). Final cumulative mortality is 6.8% in panel (a) (overshooting the graph), 0.8% in (b), 0.18% in (c), and 0.13% in (d).

## Discussion

In this study we explored how interactions between the dynamics of COVID-19 spread, policy responses to this in the form of NPI, and variations in the compliance of the population to NPI, might affect the course of the pandemie. Our model takes into consideration that policy responses are the result of complex deliberative processes that may not always be perfectly timed, and that the effectiveness of individual NPI, both at the time they are introduced and while they are maintained, can be difficult to predict. For the example of the UK we found that, as a result of these uncertainties only, there is considerable variability in the potential development of the COVID-19 outbreak and the number of lives it could take. In view of this inherent variability, it should not be a surprise that COVID-19 outbreaks in different countries are taking quite different forms, even when their governments and societies are working in similar ways.

Another surprising outcome of the present model is that, within limits, any decline of the population’s compliance through time has a comparatively weak effect on cumulative mortality: a three-fold increase in the rate of compliance decline led to less than a doubling of the median number of fatalities in the model. The reason for this is that in our model governments can counteract declining compliance by introducing new regulatory measures once the number of infections raises again. In practice this might simply involve reinforcing messages about the importance of compliance and/or increasing enforcement of the existing measures: during previous influenza pandemics declines in compliance have coincided with reductions in reported cases and deaths and associated media coverage (Lau *et al*. 2003; Wong & Sam 2010). This result is reassuring, given the scarcity of systematic knowledge about how compliance with NPI changes through the course of an epidemic.

By contrast, the frequency at which containment policies are reviewed and, if required, additional ones introduced, can have dramatic impacts on outcomes. Reduction of this frequency by a factor of two led to a more than ten-fold increase in the median number of fatalities in our model. Most of this effect arises during the initial, main outbreak (Fig. 4), but later, secondary outbreaks can also become larger as a result (e.g. Fig. 4b). The reason for this dependence is clearly the fast, exponential spread of COVID-19. To be effective, the frequency of policy revisions should be comparable to the doubling time of the outbreak. We note in this context that, with a doubling time of around 4 days, our model parameterization might be rather conservative.

Compared to other aspects of disease progression, the range of values over which our measure for the effect of NPI, *d*_*t*_, fluctuates turns out to be robust amongst simulation runs. The reason is that in most cases the majority of the population remains susceptible to the disease and the number of infected people neither increases nor decreases in the long term. The NPI in place must therefore remain such that the reproduction number remains close to 1 on average, even if it fluctuates in the short term.

The concept of management strategy evaluations originated in fisheries management (Bunnefeld *et al*. 2011; Punt *et al*. 2016), a task that has some similarity with the management of diseases. In both cases the information available about the biological system to be managed is incomplete, there is substantial inherent randomness, and yet the biological process itself is in principle well understood. Uncertainty about management outcomes therefore depends to a large part on the nature of management measures and the logic according to which they are applied. Management strategy evaluations, i.e. the modelling of the interactions between biological system and management, can then be useful for comparing the long-term implications of different management strategies.

The strategies considered in this study are only a small subset of the conceivable approaches to managing the COVID-19 pandemic. Notably, we did not include the possibility of contact tracing, which can reduce the need for social distancing measures when the number of infected individuals is low, although the resources required to do this during a pandemic make it unlikely to be used on a large scale (Wu *et al*. 2006). On the other hand, our model did not consider the uncertainty decision makers face regarding the actual number of infected individuals at a given time. Our simulation code, which we provide alongside this paper, is easily modified to model the long-term implications of these and other considerations. We invite planners to adapt it to investigate the specific questions they face.

## Data Availability

The simulation code underlying this study is available following the link below.

https://github.com/pipapu/Covid-19-Policy-Simulator/releases/tag/v1.0.0

## Acknowledgements

We thank Richard A. Nichols for insightful comments on earlier versions of the paper. This research was partially supported by the Natural Environment Research Council (NE/T003510/1).

## Code availability

Our simulation code is available at: https://github.com/pipapu/Covid-19-Policy-Simulator/releases/tag/v1.0.0

